# Immunohistochemical Study of Histone Protein 3 Modification in Pediatric Osteosarcoma Identifies Reduced H3K27me3 as a Marker of Poor Treatment Response

**DOI:** 10.1101/2023.09.13.23295510

**Authors:** Sebastian Kondratowski, Danielle Cohen, Rebecca J. Deyell, Ash Sandhu, Jonathan W Bush

## Abstract

The most common pediatric primary malignant bone tumor, osteosarcoma, is often described as genetically non-recurrent and heterogeneous. Neoadjuvant chemotherapy is typically followed by resection and assessment of treatment response, which helps inform prognosis. Identifying biomarkers that may impact chemotherapy response and survival could aid in upfront risk stratification and identify patients in highest need of innovative therapies for future clinical trials. Relative to conventional genetics, little is known about osteosarcoma epigenetics. We aimed to characterize the methylation and phosphorylation status in osteosarcoma using histone markers found in primary diagnostic biopsies and their paired metastases. We constructed two tissue microarray sets from 58 primary diagnostic samples and 54 temporally-separated but related metastatic or recurrent samples, with tissue blocks available from 2002-2022. Clinical charts were reviewed for post-therapy necrosis response, presence of metastatic disease or recurrence, and overall survival. We evaluated 6 histone H3 residues using immunohistochemistry, including H3K4me3, H3K9me3, H3K27me2, H3K27me3, H3S10T11phos, and H3S28phos. Tumors were scored with low (<25%) or high (≥25%) nuclear staining of tumor cells. Diagnostic biopsies with low H3K27me3 nuclear staining were associated with poor treatment response (≤90% necrosis) at the time of definitive excision (*P*<0.05). We observed loss of H3S10T11phos expression in metastatic and recurrent resections specimens compared to the primary tumor (*P*<0.05). Expression patterns for the remaining histone markers did not show significant associations with disease parameters or survival. Although larger cohort studies are needed, these results support the expanded evaluation of histone markers, particularly H3K27me3 and H3S10T11phos, in osteosarcoma biology and risk stratification.

## 1. Introduction

Osteosarcoma (OS) is the most common primary malignant bone tumor in the pediatric age group, and has a bimodal distribution with a second peak in older individuals [1]. Specifically within the pediatric and young adult population, OS peaks in the teenage years [2]. In pediatric and young adults <24 years of age, the incidence ranges from 2-5 per million per year in the USA [3]. Clinical presentation related to primary tumor sites may include pain, swelling or mass, constitutional symptoms and/or pathologic fracture. Historically, OS has been treated after an initial diagnostic biopsy with a standard chemotherapy protocol (MAP; methotrexate, doxorubicin, and cisplatin), and the 5-year survival rate has remained suboptimal compared to other pediatric cancers, with survival being 60-70% for those with localized disease [4]. Unfortunately, up to 20% of patients may have metastatic disease the time of diagnosis, for which 5-year survival outcomes remains dismal at <30% [5][6].

Classification of OS is typically made according to the World Health Organization (WHO), which identifies different types of OS based on location, radiology, and pathologic findings [7]. Most OS are considered conventional high grade tumors that are intra-osseus and often break through the cortex, alter the periosteum, and invade into the adjacent soft tissues. Conventional OS can demonstrate different morphologic appearances and matrix components, and while many pathologists provide a description of the morphologic appearances within the diagnosis of conventional OS, the significance of this morphologic assessment is unclear. A recent study suggested that the telangiectatic and unspecified types of conventional OS were favorable, while the chondroblastic subtype was less favorable [2].

The development and progression of OS involves genetic and molecular changes that are still not fully understood. The classic description for the underlying chromosomal instability seen in OS may include chromothripsis (*thripsis: destruction into small parts*). Chromothripsis is quite different from the step-wise fashion of oncogenesis seen in other conventional tumor progression pathways which typically involves successive mutations in proto-oncogenes or tumor suppressor genes. Chromothripsis was identified in 77% of OS, with 61% of these showing the complex and haphazard copy number variations across at least 5 chromosomes [8]. Narrowing down to individual genes, two genes are well known to be associated with OS. Classic syndromes with germline mutations leading to OS include Li-Fraumeni (*TP53* mutation; mutations seen in up to 95% of OS [9]) and hereditary retinoblastoma (*RB1* mutation; mutations seen in up to 64% of OS [10]). Interestingly, a recent study found that in fact up to one-quarter of 1,244 unselected OS patients demonstrated an underlying germline cancer predisposition variant, including genes not previously linked to OS [11]. A recent review looking at the molecular mechanisms in OS incidence and progression highlights a number of genes and cellular mechanisms that may be critical and oncogenesis and potentially therapeutically targetable, but limited work has been done in the epigenetics of OS [12].

Epigenetics involves complex processes of regulating gene expression and activity without directly altering the DNA sequence and may occur in a different pattern from cell to cell and tissue to tissue. This regulation may include post-translational histone modifications (PTHM) which allows for a particular gene to be expressed or silenced without an underlying change to the DNA sequence. Histones are proteins that are bundled in DNA to form nucleosomes, a core structure of chromatin, resulting in a ‘beads on a string’ structure. The N-terminal tails that protrude from these proteins can be remodeled with covalently attached residues such as methyl or phosphoryl groups [13]. Between these residues, molecular interactions configure euchromatin, where the DNA is readable and in the ‘beads on a string’ form, and heterochromatin, where DNA is unreadable and in the ‘30-nm fiber’ form. These chromatin remodeling mechanisms give modulatory control of gene expression within a cell, by modifying access for RNA polymerases to transcribe DNA. Despite the classical description in oncogenesis as a group of diseases born of genetic aberration, epigenetic perturbations have been found to underlie several malignancies.

The value of histone research translating to diagnostic pathology has been recently demonstrated, notably in cases of malignant peripheral nerve sheath tumors (MPNST). Cleven et al., showed that the diagnostic accuracy for MPNSTs arising from a neurofibroma could be improved by an immunohistochemistry (IHC) stain for the tri-methylation of the 27th residue (lysine) on the histone protein 3 (H3K27me3) [14]. Other studies have found that other methylation patterns in MPNST, such as dimethyl loss at H3K27, may be more specific when using immunohistochemistry [15]. Since its role in MPNST, H3K27me3 has been implicated in a number of different tumors for diagnostic and prognostic purpose, often associated with a poor prognosis or aggressive behavior, including rhabdomyosarcoma [16], chordoma [17], CNS tumors [18], amongst others.

There are other markers of post-translational modification of H3, which are generally associated with DNA replication in the S-phase of the cell cycle [19]. A select number of these residue alterations have been reported in the literature (Table 1). The function of these specific PTHMs are usually context-specific but have been studied in depth for their abilities to form areas of closed or open chromatin at varying times in development, throughout the cell cycle, or by modulating the actions of other epigenetic factors. Therefore, these dynamic controlling mechanisms make PTHMs particularly interesting as developmental or cell cycle dysregulation are both hallmarks of malignancies.

**Table 1.** An overview of each of the PTHM in this study and their known epigenetic functions and utility.

| Post-Translational Histone Modification(s) | Shorthand | Epigenetic Function/Information | Studied in Pediatric Cancer Type |
| --- | --- | --- | --- |
| Tri-methylation of the 4th residue (lysine) on histone protein 3 | H3K4me3 | Upregulates neighboring genes by forming areas of open chromatin. Involved in stem cell potency and cell differentiation and is ubiquitous in regions of DNA associated with development [37]. | Ewing Sarcoma [38], Rhabdomyosarcoma [39]. |
| Tri-methylation of the 9th residue (lysine) on histone protein 3 | H3K9me3 | Forms areas of closed chromatin. Imperative to development, embryogenesis, and cell lineage commitment [40]. | Gliomas [41][42]. |
| Di-methylation of the 27th residue (lysine) on histone protein 3 | H3K27me2 | Ubiquitous PTHM that is postulated to exert control over genetic enhancer regions that modulate non-cell-specific genes. Expression of this marker is hypothesized to prohibit acetylation of lysine 27 residues on H3 [43]. | Malignant Peripheral Nerve Sheath Tumor (MPNST) [44]. |
| Tri-methylation of the 27th residue (lysine) on histone protein 3 | H3K27me3 | Downregulates neighboring genes by forming areas of closed chromatin [14][43]. | Ependymomas [45], MPNST[46], Rhabdomyosarcoma [16]. |
| Phosphorylation of the 10th residue (serine) and 11th residue (threonine) on histone protein 3 | H3S10T11 phos | This combinatorial PTHM has been shown to enhance the promoters of cell cycle regulators, increase H3K4me3 acetylation, and therefore transcriptional activity [46][33][47]. | Neuroblastoma [48]. |
| Phosphorylation of the 28th residue (serine) on histone protein 3 | H3S28phos | Context specific. Forms regions of closed chromatin during mitotic and meiotic events, and open chromatin for transcriptional activities [49]. | Osteosarcoma [50]. |

In this current study, our primary aim was to survey selected epigenetic markers in a series of diagnostic OS biopsies to determine if any particular patterns of open or closed chromatin may predict neoadjuvant necrosis response, overall survival and other important clinical outcomes such as posttherapy response and event free survival. Our secondary aim was to determine if there were any specific changes seen when the diagnostic biopsy was compared to a matched metastatic or recurrent biopsy.

## 2. Materials and Methods

### 2.1 Sample Identification

This project was approved by the British Columbia (BC) Children’s and Women’s Research Ethics Board. The pathology laboratory information system was queried using the term “osteosarcoma” and “osteogenic sarcoma” between 2002 and 2022 which identified primary diagnostic biopsies and metastatic and recurrent specimens. Clinical charts were reviewed when available to collect demographical data, disease characteristics and investigations, therapy, and disease events, last known medical follow up, and survival status is known. Cases were excluded from analysis if the formalin-fixed, paraffin-embedded (FFPE) block(s) were not available.

### 2.2 Tissue Microarray and Immunohistochemistry

Tissue microarrays (TMAs) were built using the Manual Tissue Arrayer (Beecher Instruments Incorporated, Sun Prairie, WI). Hematoxylin and eosin (H&E) slides from the available FFPE blocks were reviewed and areas selected for TMA inclusion were selected that were most representative of the tumor and with no necrosis. In samples that showed a variety of morphologies, areas of morphologic heterogeneity were intentionally selected. TMAs were constructed with duplicate 1.0 mm cores for each case in each TMA, and two TMAs were constructed for the primary and event tumors each (four total TMAs). Non-malignant tissues were interspersed in the TMA, including normal cortical bone, bone marrow, osteochondroma, chondroblastoma, and osteoblastoma. After reviewing the TMA H&E slides, the TMAs were stained using the following IHC antibodies: H3K4me3 (Abcam, ab8580; 1:1000), H3K9me3 (Abcam, ab176916; 1:500), H3K27me2 (Abcam, ab24684; 1:200) H3K27me3 (Cell Signaling Technology, C36B11 9733; 1:200), H3S28phos (Abcam, ab32388; 1:10,000), and H3S10T11phos (Abcam, ab32107; 1:100). Antibodies were optimized using the known positive and negative controls listed by the manufacturer.

Scoring and interpretation of the TMAs was done using QuPath [20] under the guidance of the study pathologist (JWB). Relative positivity values used in the comparison of primary tumors and their paired first events (metastasis or recurrence) were calculated using the positive cell detection function. We used the highest interpretable cores to analyze our primary as well as their paired first event tumors. This follows the default scoring system for many studies involving TMAs [21]. Staining in known normal tissues were excluded from analysis. Each antibody was tested against two cores from each tumor block to address tumor heterogeneity. The scoring system used a dichotomous scale of low (<25%) and high (≥25%) tumor nuclei expression. Dropped cores from the TMA or cores that did not include tumor were excluded from analysis.

### 2.3 Statistical Analysis

Outcomes including post-chemotherapeutic response at the time of definitive resection, represented by tumor necrosis percentage, were reformatted to binary values, being either a good (≥90% necrosis) or poor (<90% necrosis) response to treatment. TMA scoring categories were also formatted to binary values of low expression (<25%) and high expression (≥25%). Pairwise deletion was used in these cases of missing data, whether it be unavailable due to missing information in the clinical charts, or randomly lost data (i.e. dropped TMA cores). Data were summarized through descriptive statistics including frequencies and percentages for categorical variables. ***X***^2^ contingency analyses were performed and p-values of <0.05 were deemed to be statistically significant for this study. Fisher’s exact test was utilized in cases where the expected counts violated the assumptions of the analysis. The relationship between survival and histone modification was summarized Kaplan-Meier survival curves. Statistical analysis was performed using R statistical software (R Core team 2023, Vienna, Austria).

## 3. Results

### 3.1 Patient Information

All patients were aged 19 or younger at the time of their surgical procedure. There were 58 available diagnostic OS biopsies identified and 54 metastatic or recurrent samples from 20 of the original 58 diagnostic biopsy patients. Amongst those 20 patients with metastatic or recurrent tumors, the median number of samples per patient was 1.5 (range of 1 to 5). Overall survival was determined by ‘enrolment’ at the date of diagnosis up until either an event or the last known healthcare follow-up (censored). Among 58 included patients, 60% had localized disease only, 57% were male, and the median age at primary diagnosis was 13.83 years old (range of 4.43 to 17.84 years old), similar to published literature (Table 2) [22]. The histologic morphologies seen in our patient cohort also follow the expected distribution with the osteoblastic morphology (Fig. 1A,B) being the most common in 70.6% of patients [23]. Lastly, the outcomes in our patient cohort is consistent with literature, where we found that nearly 70% of patients had a favorable 5-year overall survival and patients with only localized disease had superior survival outcomes (p=0.0058)[12].

**Table 2.** An overview of the study’s patient demographics (n=58).

| Category | Counts | Percentage |
| --- | --- | --- |
| **Sex** |  |  |
| Male | 33 | 57% |
| Female | 25 | 43% |
| **Age at Diagnosis** |  |  |
| 0-9 years old | 8 | 14% |
| 10-14 years old | 36 | 62% |
| 15 years old and over | 14 | 24% |
| **Histological Variant** |  |  |
| Osteoblastic | 36 | 62% |
| Fibroblastic | 2 | 3% |
| Chondroblastic | 5 | 9% |
| Telangiectatic | 5 | 9% |
| Other (parosteal, periosteal, small cell, hybrid) | 10 | 17% |
| **Tumor Size (by greatest dimension at resection)** |  |  |
| ≤ 8 cm | 21 | 36% |
| > 8 cm | 23 | 40% |
| Unknown | 14 | 24% |
| **Pathology Status** |  |  |
| Localized Only | 35 | 60% |
| Metastatic | 16 | 28% |
| Recurrence at primary site | 4 | 7% |
| Unknown | 3 | 5% |
| **Response to Neoadjuvant Therapy** |  |  |
| Good (≥ 90% tumor necrosis) | 24 | 41% |
| Poor (< 90% tumor necrosis) | 28 | 48% |
| Unknown | 6 | 10% |
| **Overall Survival** |  |  |
| Alive | 36 | 62% |
| Deceased | 21 | 36% |
| Unknown | 1 | 2% |

**Figure 1.**
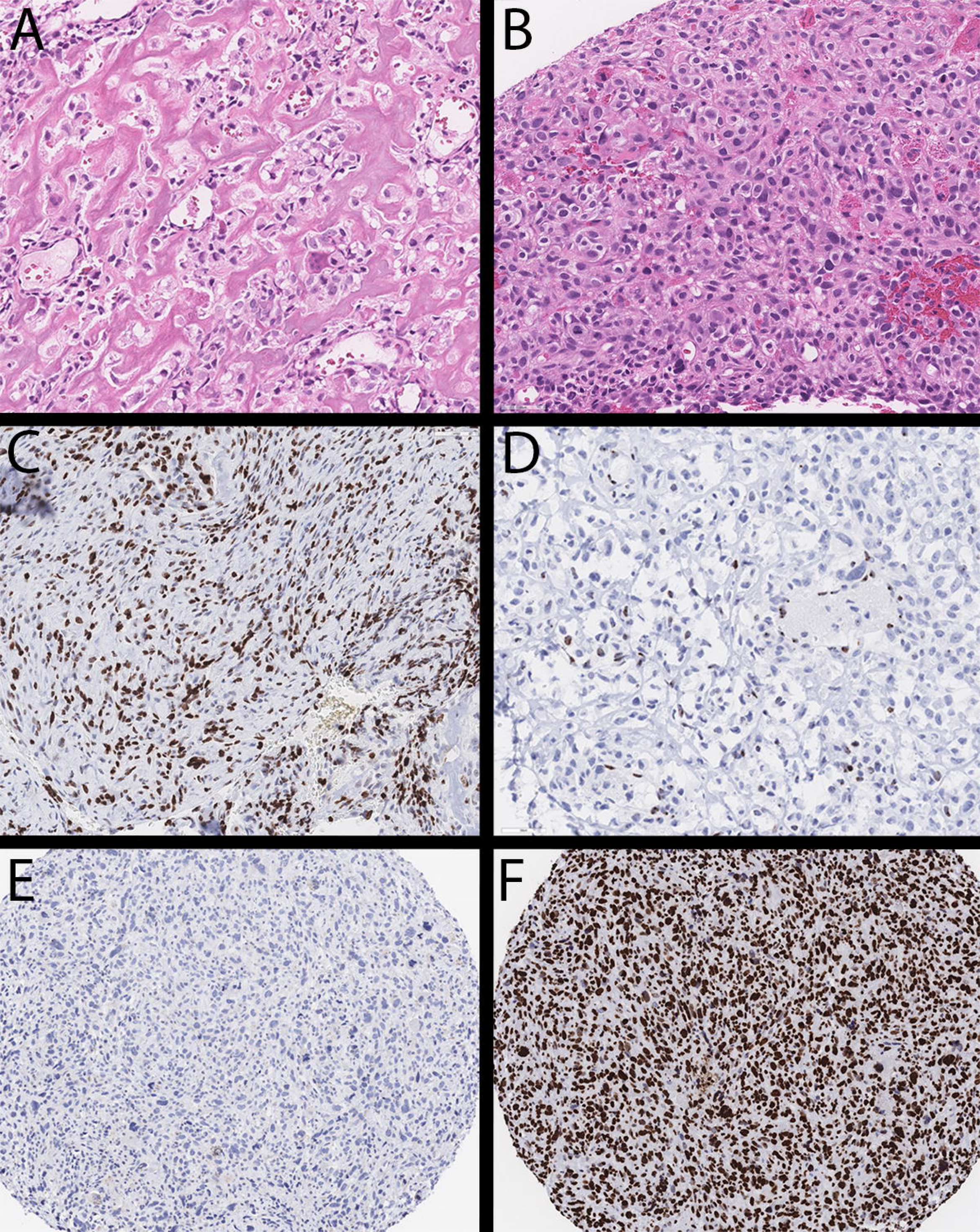
Photomicrograph showing typical morphology in conventional high-grade osteosarcoma (A,B; hematoxylin and eosin; 40x magnification). H3K27me3 immunostaining considered high/retained (C; 40x magnification) and low/loss (D; 40x magnification). All samples showed diffuse loss for nuclear H3K4me3 staining (E; 20x magnification), while all samples showed diffuse positivity in tumor nuclei for H3K9me3 (F; 20x magnification).

### 3.2 Loss of H3K27me3 Staining is Significant in Diagnostic Tumors

Amongst the samples in the diagnostic tumor TMA, we found that loss of H3K27me3 was associated with a poor neoadjuvant response (p=0.0224) (Fig. 1C, D). Of the 26 poor responders, 22 had loss of the H3K27me3 nuclear stain within tumor cells in diagnostic biopsies. However, the pattern of H3K27me3 was not predictive of overall survival (Fig. 2A), or other clinical outcomes.

**Figure 2.**
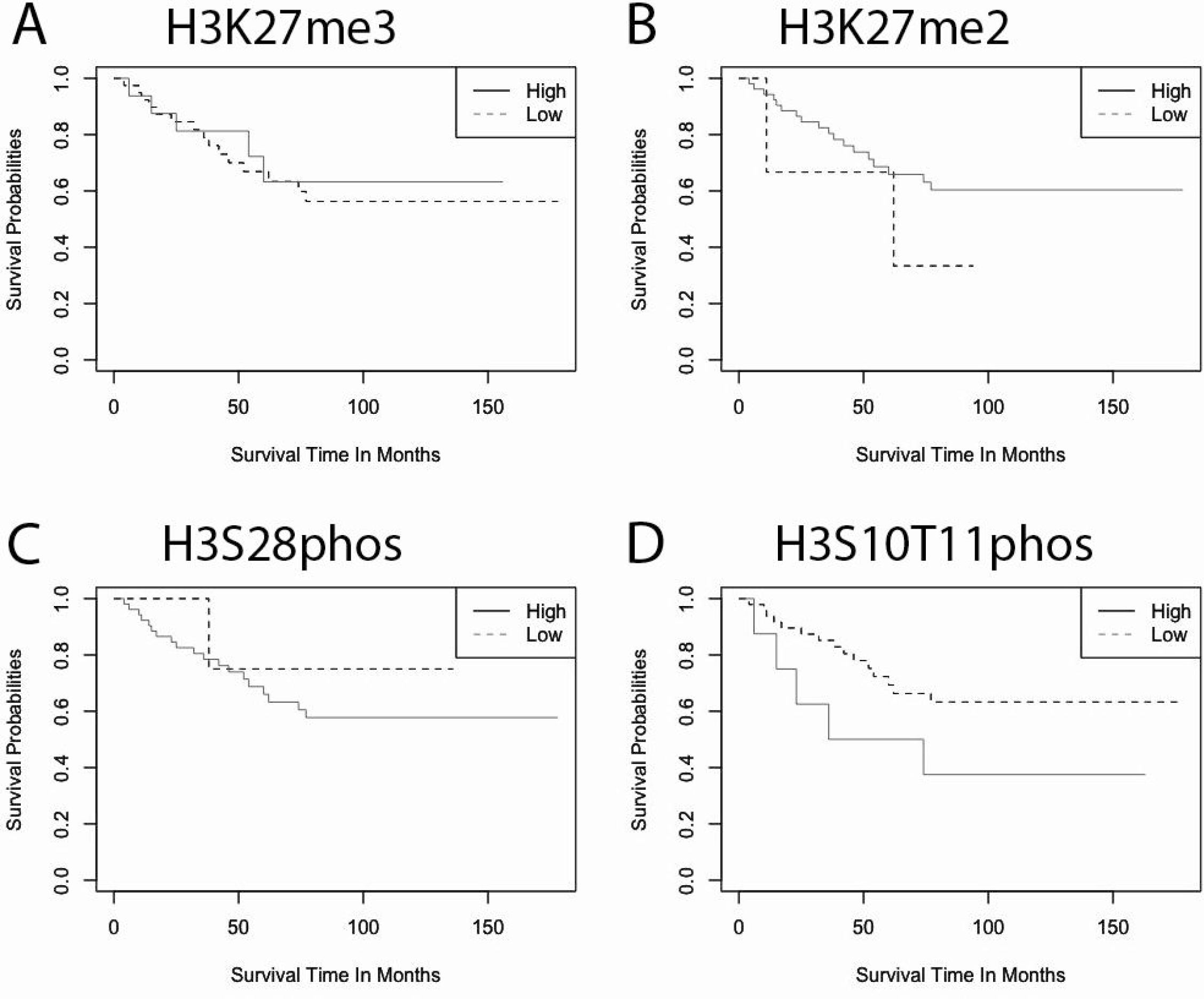
Kaplan-Meier curves that compare the survival outcomes of patients that had either retention (≥25% of tumor nuclei stained) or loss (<25% of tumor nuclei stained) for the given PTHMs (A: H3K27me3; B: H3K27me2; C: H3S28phos; D: H3S10T11phos).

### 3.3 Loss of H3K4me3 and Retention of H3K9me3 Nuclear Staining Observed in All Primary and Subsequent Event Tumor Nuclei

Interestingly, H3K4me3 immunoexpression was found to be low to nearly absent (<25% of tumor cells) in all tumor cores upon TMA analysis (Fig. 1E). On the contrary, H3K9me3 nuclear expression (≥25% of tumor cells) was observed in almost all TMA cores at the time of analysis (Fig. 1F). We found that these patterns remained consistent in the paired metastases and recurrences on our metastatic and recurrent TMA sets.

### 3.4 H3K27me2 and H3S28phos Immunoexpression is not Associated with Measured Clinical Outcomes in Primary Cases

Observed counts of H3K27me2 and H3S28phos were not predictive of survival outcomes, post-therapy response, or EFS in our cohort. (Fig. 2B, C).

### 3.5 First Event Tumors Have Near Complete Reduction of H3S10T11phos Expression Relative to Their Primary Tumors

In comparing the expression of H3S10T11phos between primary and paired metastases and recurrence, we found that the latter have significantly lower expression (p=0.011) (Fig. 3A). We noted other interesting patterns, such as reduced expression in H3K27me3 compared to primary tumors (Fig. 3B), but these results were not significant. Despite this pattern in comparing primary and first event tumors, H3S10T11phos expression on diagnostic biopsy was not found to be predictive of overall survival or necrosis response. Although not statistically significant (p=0.096), a trend was seen for a favorable prognosis when expression levels were reduced (Fig. 2D). The results of global PTHM changes were not significant for H3K27me2 (Fig. 3C) or H3S28phos (Fig. 3D).

**Figure 3.**
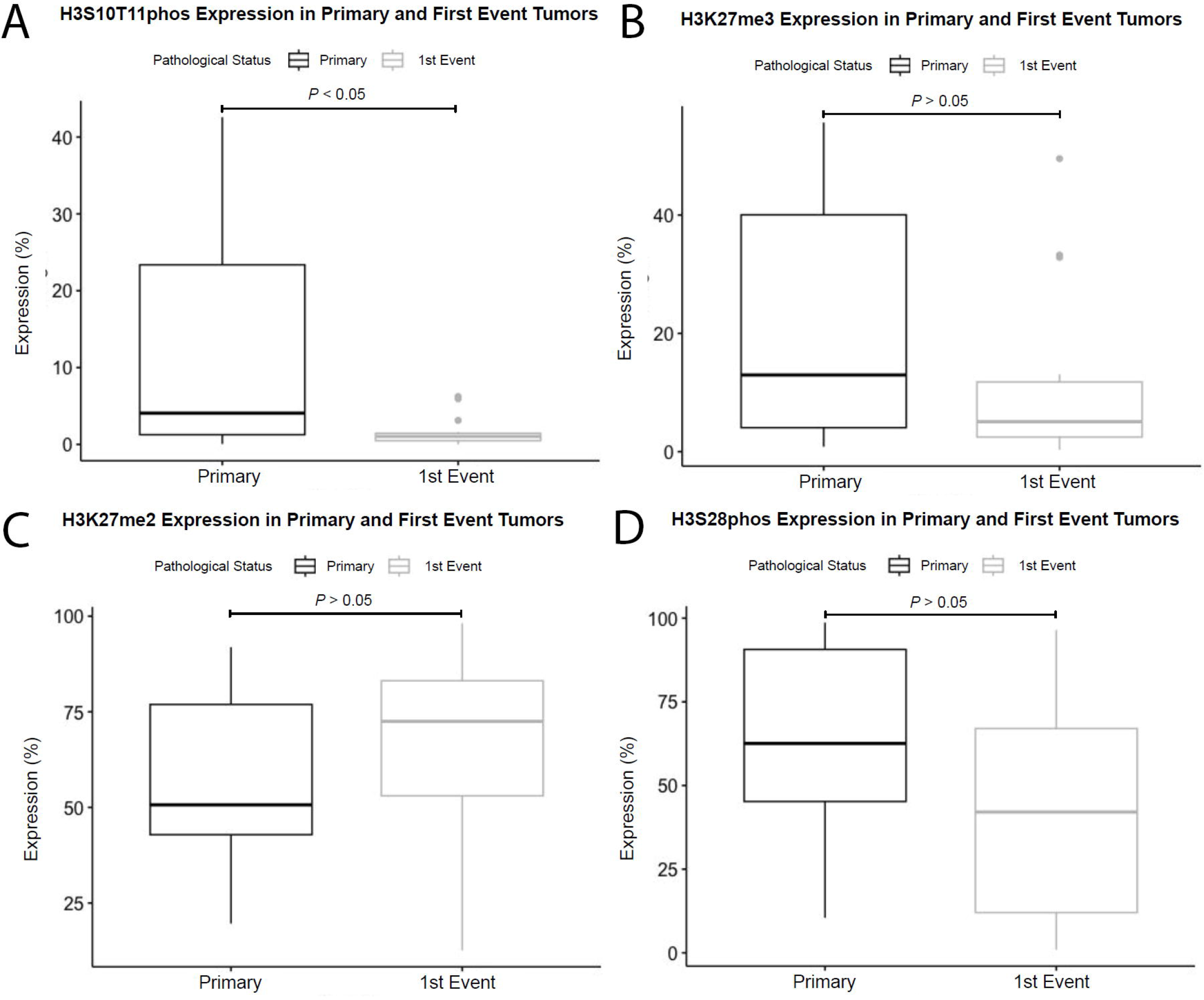
Differences in expression of post-translational histone modifications between pre-therapy diagnostic biopsies and subsequent paired first events. A: H3S10T11phos demonstrated a notable decline in expression in the first event tumor (mean difference: 10.74; 95% C.I. 2.85 - 18.63; p<0.05). There were no significant relationships found between primary and first event tumors for H3K27me3 (B), H3K27me2 (C), or H3S28phos (D).

## 4. Discussion

We report the first study that provides an immunohistochemical survey of epigenetic H3 alterations in diagnostic OS biopsies, with an enhanced analysis by evaluating the changes seen in available paired metastases or recurrences. In our study, we found that reduced expression of H3K27me3 is associated with poor response to neoadjuvant therapy at the time of definitive resection. Previous research has demonstrated a relationship between cisplatin and H3K27me3, suggesting that resistance to cisplatin was inversely related to H3K27me3 expression in *in vitro* and *in vivo* models [24]. Our results corroborate these findings as the loss of H3K27me3 may predict poor response to neoadjuvant therapy, which routinely includes cisplatin. Therefore, further studies may seek to explore the possibility of including more intensive or novel therapies at the outset of treatment for localized OS patients who demonstrate loss of H3K27me3 on their diagnostic biopsies. Limited studies elsewhere have explored H3K27me3 in OS. One of these few studies found that 3 of 19 OS cases demonstrated loss of this marker by IHC, and did not show molecular or other features to suggest that these H3K27me3-negative OS cases were not MPNST with heterologous elements [25]. The remaining studies of H3K27me3 in OS generally focus on therapeutic considerations, such as the role of methionine metabolism [26].

Another relationship we identified was that H3S10T11phos exhibits low or absent IHC expression at the time of first relapse, often when positive in the preceding primary biopsy. This phenomenon is interesting as there was no statistically significant difference in survival for those with low or high expression of H3S10T11phos at the time of diagnostic biopsy. The expression of H3S10 phosphorylation has been shown to be associated with a more aggressive form of gastric cancer with loco-regional recurrence [27], and it is believe that the STAT3 and mitogen-and stress-activated protein kinase 1 (MSK1) are involved in phosphorylating H3S10 in these aggressive forms of gastric cancer [28]. STAT3 has been implicated in a number of cancers, including OS [29], where overexpression is often seen in cases with worse outcomes, with an exception being breast cancer. MSK1 is part of the MAPK pathway, being regulated by p38 MAPK [30]. However, the role of MSK1 is not well understood in OS in the context of overall MAPK/ERK pathway [31]. Evaluation of these two H3S10 phosphorylation regulators may be high-value targets, biologically and therapeutically, to understand our observation of marked reduction in H3S10T11phos IHC expression in relapse specimens. Comparing pathway alterations at the transcriptome or proteome level between diagnostic and recurrence biopsies may further our understanding of developing metastatic potential in OS. H3S10phos has been observed to be co-expressed with H3K9me3, a marker of heterochromatin, and downregulates it prior to mitosis likely to allow for DNA replication [32]. This is quite interesting, as we found that all cases that subsequently lost H3S10T11phos retained their H3K9me3 staining. On the other hand, H3T11phos has been shown to be a marker that optimizes demethylation of tri-methylated residues [33]. For example, H3K27me3 is a marker of closed chromatin, so demethylation may contribute to increased gene expression of previously closed regions. The combination of the two markers, and perhaps overexpression, may contribute to a cascade where large regions of heterochromatic DNA are made accessible. This dysregulation of gene expression could be identified as a major factor in the proliferation of OS. However, much remains to be discussed, which cannot be done without more precise techniques that can highlight which regions of DNA are being affected. Moreover, these results also highlight the interplay and contextual factors that occur between PTHM markers which are yet to be explored in OS. Nonetheless, this evidence provides new information on the fluidity of the epigenetic landscape of OS with significant pattern changes between primary tumors and their metastases or recurrences.

Limitations of this study are notable that this was a pilot study using a small and heterogenous cohort size of convenience. However, this method does allow for our study to identify potential candidate IHC markers of interest that may be better assessed in properly-powered studies. Given that OS samples are often hard and require decalcification, we were required to work up our antibodies under both non-decal and decal conditions. To do this, we utilized known normal and abnormal staining tissues as identified by the manufacturer, and tested these antibodies on these known tissues under both routine FFPE preparation and FFPE-decal preparations. Another limitation is that our study used a TMA-based assay, which is not able to assess for possible intra-tumoral heterogeneity. Epigenetic regulation is known have a spatial heterogeneity in many tumors [34], and this could be magnified as a source of sampling error in an OS TMA with known epigenetic and histologic heterogeneity [35][36]. Given the findings in this study, future studies may benefit from both bulk and spatially resolved analytics, including transcriptomic and proteomic studies to determine if epigenetic regulation can be used in a prognostic or predictive fashion.

## 5. Conclusions

In summary, we demonstrate that OS demonstrates intra and inter-tumoral heterogeneity by immunohistochemistry for a number of PTHM markers on diagnostic biopsies at the H3 residue. Of these, H3K27me3 shows promise as a marker of neoadjuvant response regardless of the histomorphology observed. On the other hand, we demonstrate that H3S10T11 appears to show a dramatic drop in expression level in metastatic and recurrent specimens which may indicate potential underlying biologic changes facilitate the development of metastatic or recurrent disease.

## Data Availability

Data is available upon request.

## 6. Acknowledgements

This research was supported by the Society for Pediatric Pathology Young Investigator Research Grant. Support was also provided by the BC Children’s Hospital Biobank for sample acquisition and data collection. Antibodies were optimized by Shelley Moerike, Technical Coordinator at the BCCH Anatomical Pathology Lab. Ethics and other research administration support was provided by Michelle Dittrick, Pathology Research Manager at BCCH.

## 7. Authors’ Contributions

SK: case and data acquisition, tissue microarray construction, immunohistochemistry analysis, and statistical analysis; DC: case and data acquisition, RJD: case and data acquisition, AS: statistical analysis, JWB: study design, immunohistochemistry analysis, study supervisor. All authors contributed to writing and reviewing the manuscript.

